# To pool or not to pool samples for Sexually Transmitted Infections detection in Men who have Sex with Men? An evaluation of a new pooling method using the GeneXpert instrument in West-Africa

**DOI:** 10.1101/2020.01.22.20017392

**Authors:** Irith De Baetselier, Bea Vuylsteke, Issifou Yaya, Anoumou Dagnra, Souba Diandé, Jeff Yaka, Gérard Kadanga, Issa Traore, Vicky Cuylaerts, Hilde Smet, Elias Dah, Ephrem Mensah, Camille Anoma, Amadou Koné, Diallo Dramane, Hortense Fayé-Ketté, Alain Yeo, Bintou Dembélé Keita, Christian Laurent, Tania Crucitti, for the CohMSM-PrEP study group

**Affiliations:** Institute of Tropical Medicine, Department of Clinical Sciences, Antwerp, Belgium; Institute of Tropical Medicine, Department of Public Health, Antwerp, Belgium; IRD, Inserm, Univ Montpellier, TransVIHMI, Montpellier, France; CHU-SO-LNR-TB, Lomé, Togo; LNR-TB, Ouagadougou, Burkina Faso; Espoir Vie Togo, Lomé, Togo; Association African Solidarité, Ouagadougou, Burkina Faso; Institut National de Santé Publique, Centre Muraz, Bobo Dioulasso, Burkina Faso; Espace Confiance, Abidjan, Côte d’Ivoire; SEREFO/UCRC, USTTB, Bamako, Mali; Institut Pasteur Côte d’Ivoire, Abidjan, Côte d’Ivoire; ARCAD-SIDA, Bamako, Mali; Centre Pasteur du Cameroun, Yaoundé, Cameroon

**Keywords:** Sexually transmitted infections, Africa, pooling, pre-exposure prophylaxis, men who have sex with men

## Abstract

**Background:** Men who have sex with Men (MSM) using Pre-exposure prophylaxis (PrEP) are at risk for Sexually Transmitted Infections (STIs). Therefore, PrEP services should include regular screening for *Chlamydia trachomatis* (CT) and *Neisseria gonorrhoeae* (NG) at urethra, anorectum and pharynx. However, financial and logistic challenges arise in low resource settings. We assessed a new STI sample pooling method using the GeneXpert instrument among MSM initiating PrEP in West-Africa.

**Methods:** Urine, anorectal and pharyngeal samples were pooled per individual for analysis. Unpooled samples were analyzed in case of an invalid or positive result of the pool, to identify the infection’s biological location. The results of two different pooling strategies were compared against a gold standard.

**Results:** We found a prevalence of 14.5% for chlamydia and 11.5% for gonorrhea, with a predominance of infections being extra-genital (77.6%). The majority of infections were asymptomatic (88.2%). The pooling strategy with unpooling of invalid results only, had a sensitivity, specificity and agreement for CT of 95.4%, 98.7% and 0.93, respectively; and 92.3%, 99.2% and 0.93 with additional unpooling of positive results. For NG, these figures were 88.9%, 97.7% and 0.85 for testing of invalid results, and 88.9%, 96.7% and 0.81 with unpooling of positive results.

**Conclusion:** West-African MSM have a high prevalence of extra-genital and asymptomatic STIs. The GeneXpert method provides an opportunity to move from syndromic towards etiological STI diagnosis in low income countries, as the platform is available in all African countries for tuberculosis testing. Pooling will reduce costs of triple site testing.

## Introduction

The incidence of Sexually Transmitted Infections (STIs), including *Chlamydia trachomatis* (CT) and *Neisseria gonorrhoeae* (NG), is increasing globally. Their burden is disproportionally higher in low- and middle-income countries (LMIC) and in key populations, such as men who have sex with men (MSM) [1]. Fast detection and treatment of STIs is essential, as they raise serious health concerns, including increased risk of acquiring HIV infection [2].

UNAIDS recommends a combination strategy of biomedical, behavioral, and structural approaches for HIV prevention [3]. The use of Pre-exposure prophylaxis (PrEP) is an effective new biomedical HIV prevention tool, which is increasingly used among MSM in many high-resource countries [4,5]. However, PrEP may lead to a decrease in condom use and hence enhance STIs [6]. Indeed, PrEP demonstration studies among MSM in high-resource settings reported a high STI prevalence and incidence whereby most of the STIs were of extra-genital origin i.e. pharynx and anorectum. These STIs are frequently asymptomatic [7–11].

Although frequent screening of STIs among MSM in high-resource settings is currently debated [12], African MSM often report sexual relations with women. This sexual behavior may contribute to the spread of STIs to the general population. Hence, fast detection and treatment of STI infections is recommended in this population [13].

The World Health Organization (WHO) therefore advocates the integration of STI testing and treatment in all PrEP services, so that populations at risk have access to both STI prevention and care [14]. Furthermore, triple-site testing is recommended in MSM [15,16].

To date, nucleic acid amplification tests (NAATs) are the recommended diagnostic methods to detect STIs due to their high sensitivity and specificity. However, this method requires a state-of-the-art molecular laboratory and highly trained laboratory technicians [17,18]. Unfortunately, the screening of STIs using NAATs is hampered or even absent in LMIC due to the lack of adequate laboratory services and limited resources. Because of these barriers, LMIC use a syndromic approach for the diagnosis and treatment of symptomatic STIs.

The GeneXpert platform (Cepheid, Sunnyvale, US) holds promise as a method to detect STIs in LMIC. This platform is a molecular assay which requires minimal training and yields results within two hours. Since 2010, the WHO has recommended the use of the GeneXpert platform for the confirmation of tuberculosis and the detection of rifampicin resistance of *Mycobacterium tuberculosis*. As a consequence, the GeneXpert platform has become widely available throughout Africa [19]. In 2012, the Food and Drug Administration approved a GeneXpert cartridge to simultaneously detect CT/NG in genital samples and, quite recently, to test samples of pharyngeal and anorectal origin [20]. However, the high cost of the GeneXpert CT/NG cartridge hinders its utilization for the diagnosis of CT/NG in Africa.

In addition, testing one genital (urine) and two extra-genital samples (anorectal and pharyngeal) for STI screening in MSM will further increase this cost. Hence, pooling of the three collected samples per individual offers potential cost-savings in CT/NG detection in MSM presenting for PrEP [21]. Several pooling methods are described, including two using the GeneXpert instrument for STI detection [21–25]. To our knowledge, none of these pooling methods have been implemented in LMIC. In addition, very few are able to determine the biological location of the infection, which could be important for treatment and surveillance purposes.

We evaluated the performance of a new pooling method using the GeneXpert platform for the detection of CT and NG among MSM initiating PrEP in four West-African countries.

This article was submitted to an online preprint archive [26].

## Materials and Methods

### Study setting

The CohMSM-PrEP study is being conducted in four sites in West-Africa: Ouagadougou, Burkina Faso; Lomé, Togo; Bamako, Mali and Abidjan, Côte d’Ivoire. Its aim is to assess the feasibility of PrEP among a cohort of 500 MSM, including STI prevalence. Samples for CT/NG testing were collected from all participants at their initiation visit and transported to research laboratories (SEREFO, Bamako and Institut Pasteur, Abidjan) or to national reference laboratories for tuberculosis (Laboratoire National de Recherche sur la Tuberculose-TB, Ouagadougou and CHU-SO-LNR-TB, Lomé) where the GeneXpert instrument was available (hereafter called local STI laboratory).

The study has been approved by all applicable Ethics Committees and all participants provided written informed consent.

### Quality Control

The STI reference laboratory of the Institute of Tropical Medicine (ITM) in Antwerp, Belgium, ensured the reliability and quality of the results of the local STI laboratories, including compliance with Good Clinical and Laboratory Practice Standards. This laboratory provided hands-on training in sample collection, processing, and storage at study initiation in each site. An external quality control (EQC) panel was tested at study initiation and quarterly during the study. If DNA contamination was suspected, an environmental control of the laminar flow, bench, pipettes and the surface of the GeneXpert instrument was done.

### Laboratory methods

Participants provided first-void urine and a physician took two pharyngeal and two anorectal samples (Eswab™, Copan Diagnostics, Brescia, Italy). After collection, samples were stored refrigerated (2-8°C) or frozen (-20°C) on site depending on the time of transport to the local STI laboratory (2-8°C<48h<-20°C). Transport of samples was performed under temperature monitored conditions using a cool box and cooling elements. Upon sample receipt in the local STI laboratory, one aliquot (1 mL) of urine, one anorectal and one pharyngeal Eswab™ (both randomly chosen from the duplicates) were immediately frozen (-20°C) until shipment on dry ice to ITM for reference testing. Samples for local testing were stored refrigerated (2-8°C) if analysis was performed within 72 hours of collection, otherwise, samples were stored frozen (-20°C).

#### Pooling method

At the local STI laboratory a volume of 400 µL of the three samples (urine, anorectal and pharyngeal sample) per participant was transferred in a microtube (hereafter called pool). After vortexing, one mL of the pool was transferred into the CT/NG Xpert cartridge. When the result of the pool was negative, all samples were considered negative and were not individually tested. When the pool was positive or invalid, individual samples were tested as follows: 400 µL of one sample was added to 800 µL diluted Phosphate Buffered Saline (hereafter called unpooling). After vortexing, one mL was transferred into the cartridge and analyzed. Sample processing was performed in a laminar flow cabinet.

#### Gold standard test algorithm

All duplicate samples were tested individually at ITM according to the following test algorithm in place: CT/NG was detected using the Abbott RealTi*m*e (RT) CT/NG assay (Abbott Molecular, Des Plaines, Illinois, USA) according to the manufacturer’s instructions. DNA extracts of positive samples were tested by in-house real time (RT)-PCR assays for CT and/or NG, both based on previously published primer sets [27,28]. The in-house RT-PCR for CT was able to differentiate L-type from non-L types. A sample was considered positive when positive in both the Abbott and the in-house RT-PCR. An initial positive Abbott assay result followed by a negative confirmatory NAAT result was defined as ‘not confirmed’. Inhibition according to the Abbott assay was defined as ‘inhibition’.

To exclude for sampling errors and confirm the quality of the sample, the presence of human material in the duplicate sample tested at ITM was assessed using a human Endogenous Retrovirus-3 PCR (ERV-3) on anorectal and pharyngeal samples that were solely positive on site [29].

### Identification and validation of the two pooling strategies

Two different on-site pooling strategies were evaluated. Strategy 1 consisted of triple-site pooling and testing, and unpooling only when the pooled sample result was invalid. Strategy 2 consisted of triple-site pooling and testing, and unpooling when the pooled sample result was invalid or positive for either CT or NG.

The result of the two strategies was compared with the infection status according to the gold standard. A participant was defined as not infected when his three samples were all negative according to the gold standard. A participant was defined as infected if at least one sample was positive. In the event that one or more sampling site(s) were not confirmed and the other sampling site(s) were negative, the participant infection status was defined as not confirmed.

### Statistical analysis

The sensitivity, specificity, positive predictive value, negative predictive value, with 95% confidence intervals (CI) were calculated for strategies 1 and 2, excluding inhibited and not confirmed infection status. In addition, agreement of both strategies with the gold standard test algorithm was assessed by the Cohen’s kappa statistic.

All analyses were performed in STATA V15.0.

### Cost analysis

The costs of the two different screening strategies were compared against triple-site testing using the GeneXpert. The obtained prevalence of CT/NG in this study was used to simulate the costs of STI testing in a population of 500 MSM. An invalid rate of 4% was assumed.

## Results

### Patient characteristics, test results and prevalence of CT/NG

The ITM received baseline samples from 503 CohMSM-PrEP study participants. However, since the pooling method was not performed on 6 participants’ samples, samples from 497 participants were included in the analysis.

All participants were MSM, with a median age of 24years (Inter Quartile Range: 22-28).

#### Prevalence of STIs according to the gold standard algorithm

According to the gold standard test algorithm performed at ITM, the study population had a prevalence of 14.5% CT (72/497), 11.5% NG (57/497) and 22.1% CT or NG (110/497). The anorectal site was the most common infected with CT or NG (n=76; 60.8%); followed by the urethra (n=28; 22.4%) and pharynx (n=21; 16.8%). Two participants were positive in two biological sites for CT and eleven for NG. All confirmed CT positive samples were non-L genotypes.

Of the 110 infected participants, 97 of them (88.2%) reported no symptoms of STI.

#### Test results at the study sites

Using pooled samples 131 participants were positive for CT or NG: 21 had a mixed CT/NG infection; 71 were solely CT-infected; and 39 solely NG-infected. A total of 353 participants tested negative and were not further investigated. An invalid result was obtained in 13 participants.

Using strategy 1, four additional NG infected participants were detected by unpooling the pools with an invalid result.

Using strategy 2, individual testing of the pooled samples with invalid or positive results decreased the number of infected participants with CT or NG to 128: 26 with a dual CT/NG infection, 60 with CT only and 42 with NG only.

The CT/NG results obtained on site (pooled and unpooled samples) and the results obtained at ITM are available in Figure S1.

### Test and sample quality

The study sites participated in a quarterly EQC: one NG positive sample was missed; no false positive results were reported.

The number of samples positive for CT/NG, whatever the biological site, at the study sites was systematically higher as compared to the numbers found at ITM (Fig S1). One environmental check revealed the contamination of the GeneXpert instrument’s surface with CT at one site. The contamination is probably the cause of overreporting CT in this site. Another site had a large number of falsely detected NG, however, contamination with NG was not detected during the quarterly EQC assessments and the environmental check.

The presence of human DNA was assessed in 43/46 individual extra-genital samples and in 11 (9 anorectal and 2 pharyngeal) (25.6%) of them human DNA was not detected.

### Performance of the two pooling strategies using the GeneXpert method

Samples from the CT contaminated site collected after 31/08/2018 were excluded from statistical analyses, which limited the number of participants to 448. The Supplementary Material documents all discordant cases (table S1 and table S2).

The evaluation of the two pooling strategies using the GeneXpert against the gold standard for CT and NG is presented in Tables 1 and 2.

**Table 1:**
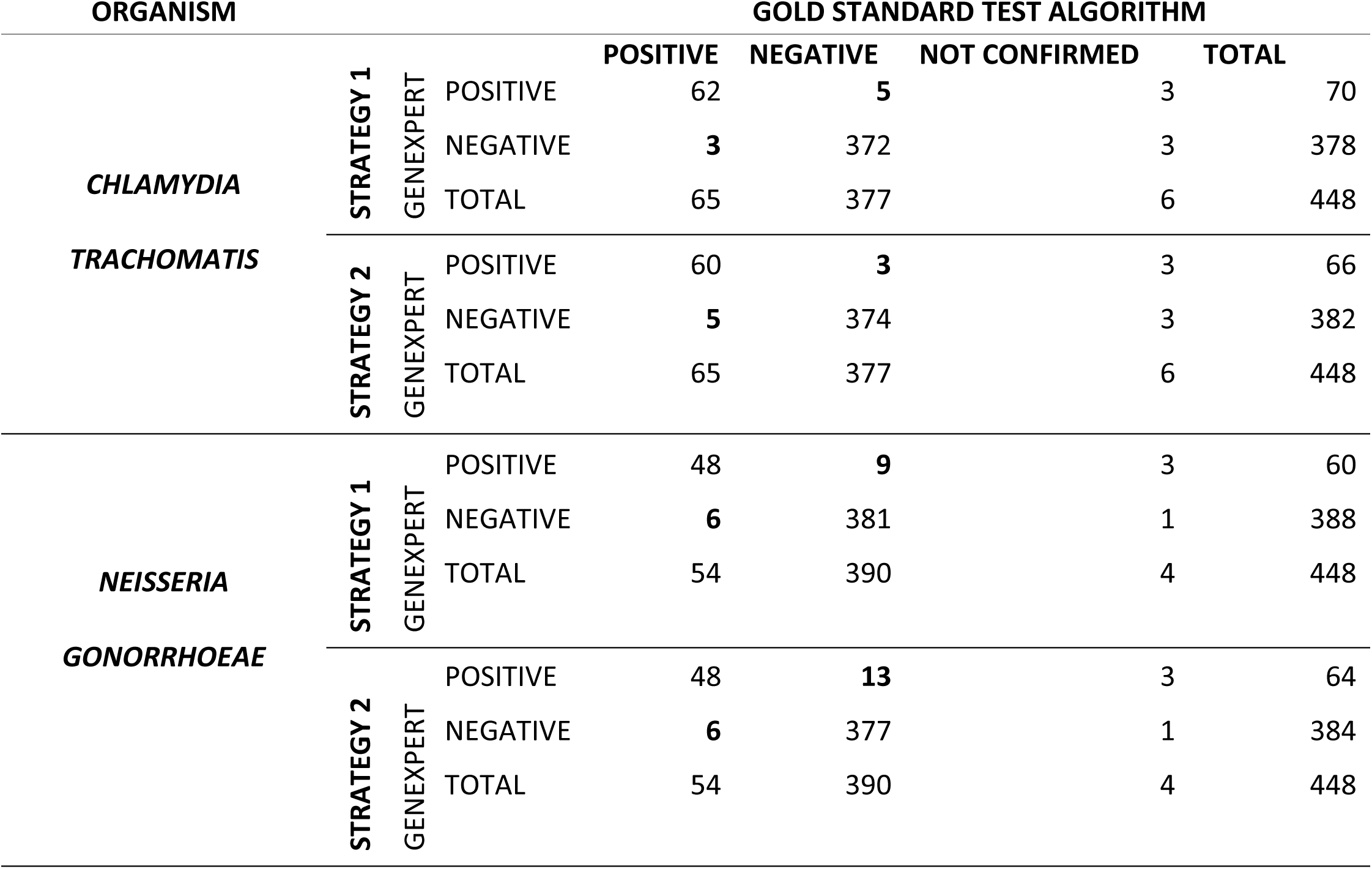
Comparison of the two test strategies to detect *Chlamydia trachomatis* and *Neisseria gonorrhoeae*. Strategy 1 will only test the samples individually when the pooled sample result was invalid. Strategy 2 will test the samples individually when the pooled sample result was invalid or positive for either CT or NG. The discordant samples are explained in detail in the Supplementary material (Table S1 and Table S2).

**Table 2:** Evaluation of the two pooling strategies against the gold standard algorithm for either *Chlamydia trachomatis* and *Neisseria gonorrhoeae*. INV: invalid; POS: positive; PPV: Positive Predictive Value; NPV: Negative Predictive Value; K-coeff: Kappa-coefficient; CI: Confidence Interval

|  |  | SENSITIVITY | SPECIFICITY | PPV | NPV | % | K- |
| --- | --- | --- | --- | --- | --- | --- | --- |
|  |  | % (95% CI) | % (95% CI) | % (95% CI) | % (95% CI) | AGREEMENT | COEFF |
| CHLAMYDIA | STRATEGY 1 | 95.4% | 98.7% | 92.5% | 99.2% | 98.2% | 0.93 |
|  | (unpooling of INV) | (87.1-99.0) | (96.9-99.6) | (83.8-96.7) | (97.6-99.7) |  |  |
|  | STRATEGY 2 | 92.3% | 99.2% | 95.2% | 98.7% | 98.2% | 0.93 |
|  | (unpooling of INV & POS) | (83.0-97.5) | (97.7-99.8) | (86.7-99.0) | (97.0-99.6) |  |  |
| GONORRHEA | STRATEGY 1 | 88.9% | 97.7% | 84.2% | 98.5% | 96.6% | 0.85 |
|  | (unpooling of INV) | (77.4-95.8) | (95.7-98.9) | (73.5-91.1) | (96.8-99.3) |  |  |
|  | STRATEGY 2 | 88.9% | 96.7% | 78.7% | 98.4% | 95.7% | 0.81 |
|  | (unpooling of INV & POS) | (77.4-95.8) | (94.4-98.2) | (66.2-86.4) | (96.7-99.3) |  |  |

#### Chlamydia trachomatis

According to strategy 1, three CT were missed, resulting in a sensitivity of 95.4%. The Abbott delta cycle (DC) values (difference in cycle numbers between the cut-off control and the sample cycle number) of the individual specimens from two discordant pools were low, which correlates with a low target concentration. CT was falsely detected on site in five participants (specificity 98.7%). Applying strategy 2, the sensitivity decreased to 92.3% and the specificity increased to 99.2%.

#### Neisseria gonorrhoeae

Six NG infections were missed with strategy 1, yielding a sensitivity of 88.9%. The Abbott DC values for the individual specimens included in the six pools indicated a high bacterial load.

Using strategy 1, nine samples were positive but not confirmed by the gold standard algorithm (specificity 97.7%). When applying the second strategy, one sample was actually negative, however, five additional tests were positive, almost all from one study site, suggesting a possible DNA contamination (specificity of 96.7%).

### Cost analysis

A prevalence of 22% CT/NG, as found in this study, was assumed. Compared to triple testing, a 56% decrease in costs was noted with strategy one and 30% with strategy two (Table 3).

**Table 3:** Number of tests required according different strategies and cost simulation using a combined prevalence of CT/NG of 22%

| STI screening strategies | Number of participants | Calculation of number of tests required | Total number of tests | Assay cost (19 USD per test) | Cost per infection |
| --- | --- | --- | --- | --- | --- |
| Triple site testing | 500 | Every site of sampling = $500 * 3$ | 1500 | 28.500 USD | 220 USD |
| Pooling strategy 1 | 500 | 500 pooled samples = 500<br>individual testing of 20 invalid results = $20 * 3$ | 560 | 10.640 USD | 97 USD |
| Pooling strategy 2 | 500 | 500 pooled samples = 500<br>individual testing of 20 invalid results = $20 * 3$<br>individual testing of 110 positive results = $110 * 3$ | 890 | 16910 USD | 154 USD |

## Discussion

We are among the first to report on the prevalence of chlamydia and gonorrhea in MSM initiating PrEP in West-Africa. The data indicate a high prevalence of chlamydia (14.5%) and gonorrhea (11.5%), mainly in asymptomatic (88.2%) individuals. These asymptomatic infections would not have been treated according to the syndromic approach, which is currently the standard of care in LMIC. In addition, 77.6% of infections were extra-genital. These findings reinforce the recommendation that STI services, including triple-site testing, should be integrated in PrEP programs in LMIC. Therefore, we aimed to implement an STI screening strategy using the GeneXpert instrument as its availability throughout Africa will facilitate STI testing. The Xpert CT/NG assay is now put forward as a potential point-of-care assay for STI detection in remote health-care settings as it is easy, robust, and has very high analytical performance [30–32]. Previous studies showed that this technology is acceptable in identifying STIs among Sub-Saharan African young women, however, to date, no study has been performed among African MSM [32–37].

We used a new pooling method with the GeneXpert platform to screen for STIs in genital and extra-genital samples among MSM. Although the pooling method was designed as such to identify the origin of infection, we showed that there is no clinical utility. First of all, we did not detect a single case of Lymphogranuloma venereum (LGV) in our study population. LGV is frequently detected in European MSM and requires a three week treatment with doxycycline versus one week in the event of a regular chlamydia infection. Secondly, all antimicrobials recommended nowadays for the treatment of gonorrhea are equally effective in the three biological sites [19,20,38]. Furthermore, the performance of the pooling method did not improve when unpooling positive samples. Nevertheless, we favor keeping the possibility to test the individual samples in case of an invalid pooled sample result to avoid additional sample collection and subsequent delay in result reporting.

Applying strategy one resulted in nine participants not receiving treatment (9/448; 2.0%), and 14 participants receiving unnecessary treatment (14/448; 3.1%). The number of false positives increased to 16 for the individually tested samples, mainly caused by a probable contamination of *Neisseria gonorrhoeae* at one of the sites. We also report on a CT DNA contamination of the GeneXpert instrument’s surface at another site. The GeneXpert method is a closed system that reduces contamination to an absolute minimum, however, due to its very low lower limit of detection, the assay is more prone to target contamination caused by the presence of genetic targets in the work environment or by sample cross contamination.

Although the GeneXpert CT/NG assay can be integrated into remote healthcare settings, this apparent risk of contamination may lead to erroneous results, which may cause emotional distress, stigma and unnecessary antibiotic pressure. Therefore, we strongly recommend that detection of CT/NG using the GeneXpert instrument is performed under the supervision of qualified laboratory personnel.

This is, to our knowledge, the first study reporting on a sample pooling method among a large number of African MSM initiating PrEP. Other pooling methods in MSM have been published, including methods using the GeneXpert assay [23–26]. Speers and colleagues evaluated a pooling method using the GeneXpert versus single site testing using the Cobas 4800 CT/NG assay and found an excellent agreement for NG and a substantial one for CT [26]. Our pooling method scored better for CT than NG and is in accordance with the findings of Sultan *et al*. who also found a decreased sensitivity of NG detection in pooled samples [25].

There are some differences between the two GeneXpert pooling studies. Speers *et al*. added the anorectal and pharyngeal swab directly to the GeneXpert Urine Specimen Kit, limiting manipulations [26]. We opted to work with simple urine containers and Eswabs™ and to prepare a separate pool to identify the biological origin of infection.

Using Eswabs™, our pooling method can tackle one of the most important global health priorities set forward by the WHO, namely the surveillance for antimicrobial resistance of NG [39]. Future research will need to show if surveillance for AMR of NG using Eswabs™ can be implemented in LMIC.

The present study does, however, have several limitations which may explain the discordances other than target contamination. Firstly, sampling errors: we determined the presence of human DNA in the pharyngeal and anorectal samples in samples that were positive solely using the GeneXpert. One quarter of them lacked human material in the duplicate sample, although physicians were trained in sample collection in order to avoid sampling errors [23]. This could also explain the false negative pools. Secondly, the Xpert CT/NG assay can detect as little as 10 NG genome copies per reaction [32]. We cannot exclude the idea that some of the positive NG results solely obtained with the GeneXpert were truly low positive results not detectable by the gold standard. Finally, samples tested with the gold standard algorithm underwent an additional freeze-thawing cycle which may have impaired the DNA in low concentration samples.

In conclusion, we showed that MSM initiating PrEP in Africa have high prevalence rates of extra-genital and asymptomatic STIs and that African countries can perform an etiological diagnosis of STIs without implementing specialized NAATs.

The availability of PrEP in LMIC is a unique opportunity to strengthen STI services in high risk populations. The momentum is now to move to efficient STI screening and to limit onward transmission. In this new PrEP-era, the WHO, ministries of health and stakeholders at a global level will need to ensure that STI management is integrated in PrEP services, and negotiations with companies to provide the tests at affordable prices are, therefore, essential.

## Data Availability

The data supporting the findings of this publication are retained at the Institute of Tropical Medicine (ITM), Antwerp and will not be made openly accessible due to ethical and privacy concerns. 
Data can however be made available after approval of a motivated and written request to the ITM at. The ITM data access committee will verify if the dataset is suitable for obtaining the study objective and assure that confidentiality and ethical requirements are in place.

## Acknowledgements

We would like to thank all the participants, study and laboratory staff of the CohMSM-PrEP study. We would also like to thank Wendy Thijs for the data entry and Margaux Balduck for additional review of the figure and tables.

